# SoundPics: visual and auditory psychophysics reveals distinct sensory profiles that differentiate schizophrenia from bipolar disorder in early psychosis across three independent Brazilian public health clinical samples

**DOI:** 10.64898/2026.01.20.26343778

**Authors:** Maria Lucia de Bustamante Simas, Aline Mendes Lacerda, Joselma Tavares Frutuoso, Isabela de Fatima Pina de Almeida, Mateus Monteiro de Gois Barros, Katherine Karla Souza da Silva, Thays Macambira da Silva, Giuliano Baltar Melo de Souza Ramos, Tamires Lima da Silva, Naianna Mocelin Ribeiro dos Santos, Amanda Almeida Rodrigues e Silva, Kayke Kauã de Siqueira

**Author notes:** Corresponding author: Name: Maria Lucia de Bustamante Simas, Address: Laboratório de Percepção Visual, LabVis-UFPE CFCH 9° Andar, Salas 34-35, Universidade Federal de Pernambuco, UFPE Cidade Universitária, 50.670-901, Recife, PE, Brasil.

## Abstract

**Objective:** Characterization of psychosis typically relies on cognitive and behavioral assessments. This study investigates whether feature-specific psychophysical experiments can detect subtle perceptual alterations in early psychosis and serve as screening tools.

**Methods:** Patients (N=120) diagnosed with schizophrenia (SCHZ, N=45), bipolar disorder (BIP, N=36), or first-episode psychosis (FEP, N=39) were recruited from public mental health facilities in Brazil and compared with age-matched healthy controls (pooled N=94). Independent psychophysical measurements were obtained within each group. The Pictorial-Size-Test (PST) assessed visual pictorial size perception (measured in degrees of visual angle). The Sound-Appreciation-Test (SAT) assessed auditory discomfort using a sensitivity-normalization procedure.

**Results:** SCHZ perceived larger pictorial sizes than controls (d=0.63, p<0.0001), and FEP showed an even larger effect (d=2.86, p<0.0001), whereas BIP did not differ from controls on PST. All three patient groups reported higher auditory discomfort than controls on SAT: SCHZ (d=1.29, p<0.0005), BIP (d=2.73, p<0.0001), and FEP (d=1.46, p<0.0003). A hierarchical algorithm (SoundPics) combining PST and SAT differentiated SCHZ from BIP with 70.8% accuracy among clinically determined cases (11.1% undetermined).

**Conclusions:** Low-cost psychophysical measurements reveal distinct sensory profiles in early psychosis: visual pictorial size enlargement in schizophrenia-spectrum disorders and pronounced auditory discomfort in bipolar disorder. These patterns are not readily perceived in standard clinical interactions but may serve as screening tools and provide diagnostic adjunctive information. External validation was performed in FEP patients clinically suspected of schizophrenia, and ongoing work is extending this validity test to FEP patients clinically suspected of bipolar disorder.

## INTRODUCTION

Schizophrenia typically emerges between ages 15 and 35, affecting cognition, work performance, quality of life, and eventually may impair productivity for life. The disorder is associated with widespread anatomical^1–7^, functional^8–12^, biochemical^13–14^, and proteomic^15^ brain alterations.

Psychosis — a core feature of schizophrenia, bipolar disorder, and other conditions — is defined in DSM-5-TR^16^ and ICD-11-R^17^ as a cluster of symptoms including hallucinations, delusions, disorganized speech, and impaired cognition. Current clinical assessments^18–22^ and neuropsychological tests^23–25^ primarily focus on cognition, memory, and behavior, with less attention to basic sensory processing.

Recurrent psychotic episodes can cause progressive harm^1–15^, making early detection and intervention a priority. In this context, sensory tests have emerged as potential supportive tools for assessing early psychosis^26–29^.

Accumulating evidence shows that visual and auditory perceptual alterations are detectable in early psychosis^26–31^. Previous work has documented such alterations in depression and schizophrenia^26–28^, and more recently in first-episode psychosis (FEP) from 2019 to 2025.

The present study builds on empirically observed sensory changes that occur during the development of psychosis^33^. We specifically designed two low-cost psychophysical tests — the Pictorial Size Test (PST)^26–28^ and the Sound Appreciation Test (SAT)^29–30^ — to measure these perceptual alterations. Alterations in size perception of visual illusions in schizophrenia are extensively reviewed elsewhere^26–29^. Indeed, visual size perception is altered in schizophrenia patients and some other mental disorders as reported over time^26–30^. Auditory discomfort in people with established neuropsychiatric symptoms is another issue^28–29^.

Here we report studies employing the same versions of PST and SAT with populations of healthy controls, FEP, schizophrenia and bipolar disorder patients in the natural settings from three independent specialized Brazilian public mental health facilities of Pernambuco, Brazil^34–39^.

The basic hypotheses are that patients with schizophrenia will present sensory responses compatible with previous literature findings and may experience pronounced auditory discomfort. Since some exploratory pilot studies with bipolar and FEP patients showed dissimilar patterns from schizophrenia, a concomitant hypothesis is that there will be distinct visual and auditory coupled patterns for each group of patients as compared to respective controls.

Verifying these hypotheses is essential to our goal of evaluating whether these tests can serve as screening tools and provide diagnostic adjunctive information in early psychosis, particularly in distinguishing schizophrenia from bipolar disorder.

## METHOD

### Participants

The present study involved 120 diagnosed patients and 94 healthy controls. Participants were between 15 and 70 years old. Thirty-nine patients formed the first-episode psychosis (FEP) group, 45 patients the schizophrenia (SCHZ) group, and 36 patients the bipolar disorder (BIP) group. Ninety-four participants formed the age-matched healthy control groups for each patient population (HC). Group characteristics are listed in Table 1.

**Table 1.**
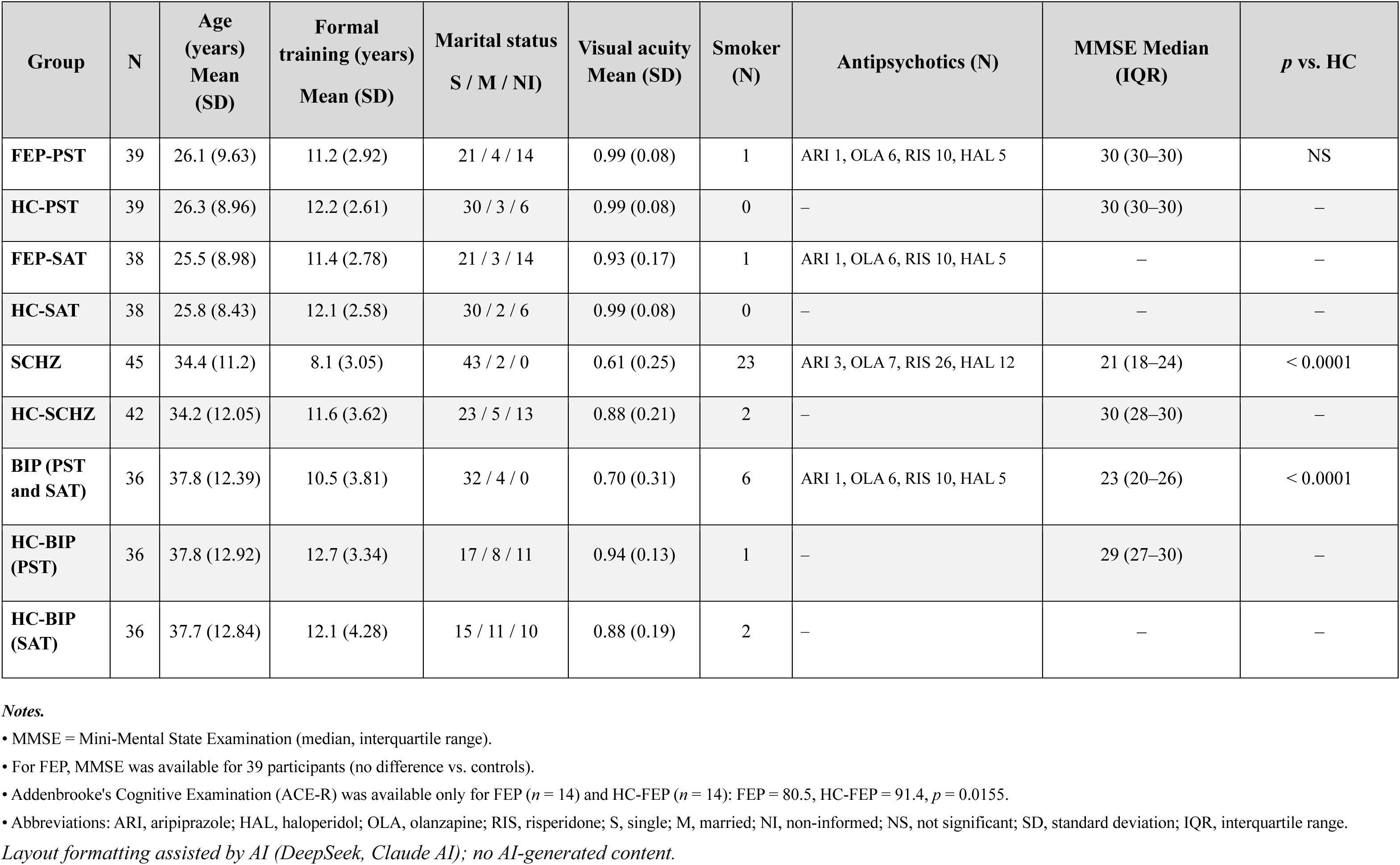
Sociodemographic and clinical characteristics of experimental and control groups.

Importantly, the three patient groups were recruited from three distinct types of mental health facilities: SCHZ predominantly from a forensic psychiatric hospital (HCTP) and community CAPS units; BIP exclusively from CAPS units in Recife and Alagoas; and FEP from a specialized first-episode psychosis outpatient clinic (AMPEP). This institutional independence supports cross-group comparisons and generalizability.

### First-episode psychosis patients

Thirty-nine participants with a provisional diagnosis of first-episode psychosis and a clinical suspicion of schizophrenia formed the FEP group. They were attending the mental health facility *Ambulatório de Primeiro Episódio Psicótico do Hospital das Clínicas da Universidade Federal de Pernambuco* (AMPEP-EBSERH-HC-UFPE). During the six-year period of the experiments, some of these patients received a definitive diagnosis and were transferred to either the SCHZ or the BIP group accordingly. Only those still under care at AMPEP remained in the FEP group sample. Not all FEP patients were medicated.

### Schizophrenia patients

The SCHZ group (N = 45) was composed of 25 males diagnosed with schizophrenia at the forensic psychiatric hospital *Hospital de Custódia e Tratamento Psiquiátrico* (HCTP) from Caetés-PE, Brazil, and 11 patients diagnosed with schizophrenia at AMPEP-EBSERH-HC-UFPE. The remaining nine patients were from outpatient CAPS (*Centros de Atenção Psicossocial*) type-II units in Alagoas (AL), Brazil. All SCHZ patients were medicated.

### Bipolar patients

The BIP group (N = 36) included 25 patients from four outpatient mental health facilities: CAPS-tipo-II *Espaço Azul*, CAPS-tipo-III *Galdino Loureto*, CAPS-tipo-III *David Capistrano*, and CAPS-tipo-III *Espaço Livremente*, all in Recife-PE, Brazil. Three patients were from CAPS-**t**ipo-II units in Alagoas (AL), and the remaining eight patients were diagnosed at AMPEP-EBSERH-HC-UFPE. All BIP patients were medicated.

### Healthy controls

Volunteers for the HC group (N = 94) were recruited from the general population; 25 of them came from the school *Educação de Jovens e Adultos* (EJA) in Arcoverde-PE, Brazil.

### Age matching

Healthy volunteers were pooled from the total of 94 to be closely paired by age during group data processing. Pairing was rigorous and produced identical mean ages with similar standard deviations in each age-matched group pair. A small subset of controls was used in more than one age-matched pair when age ranges overlapped across patient groups. This does not affect the independent comparisons, because each patient group was compared only to its own matched control set. Generally, the same HC sample served for both PST and SAT comparisons within a given patient group, and independent HC groups were composed for each age group of each patient group. There were no missing cases.

### Inclusion and exclusion criteria

Inclusion required that patients be regularly attending the mental health facility, be willing to participate, and be able to understand and follow the test instructions. There was no age restriction. Even a patient with a hearing aid was willing to participate and demonstrated adequate comprehension and task performance.

The main exclusion criterion was inability to understand instructions or agitation that precluded participation.

### Main Instrument (SounDPicS)

#### (i) The Pictorial Size Test (PST)

The present version of the PST consists of 20 color photographs of natural rural scenes (15 cm × 10 cm) bound in a 20-page folder. Ten photographs have vertical symmetry axes and ten have horizontal symmetry axes. Stimuli were viewed at a distance of 30 cm using a portable apparatus^27^ (Figure 1). Participants were instructed to circle the first image they perceived in a glimpse, after one training trial. The diameter of each circled area was measured in centimeters and converted to degrees of visual angle (dva) based on the fixed viewing distance.

**Figure 1.**
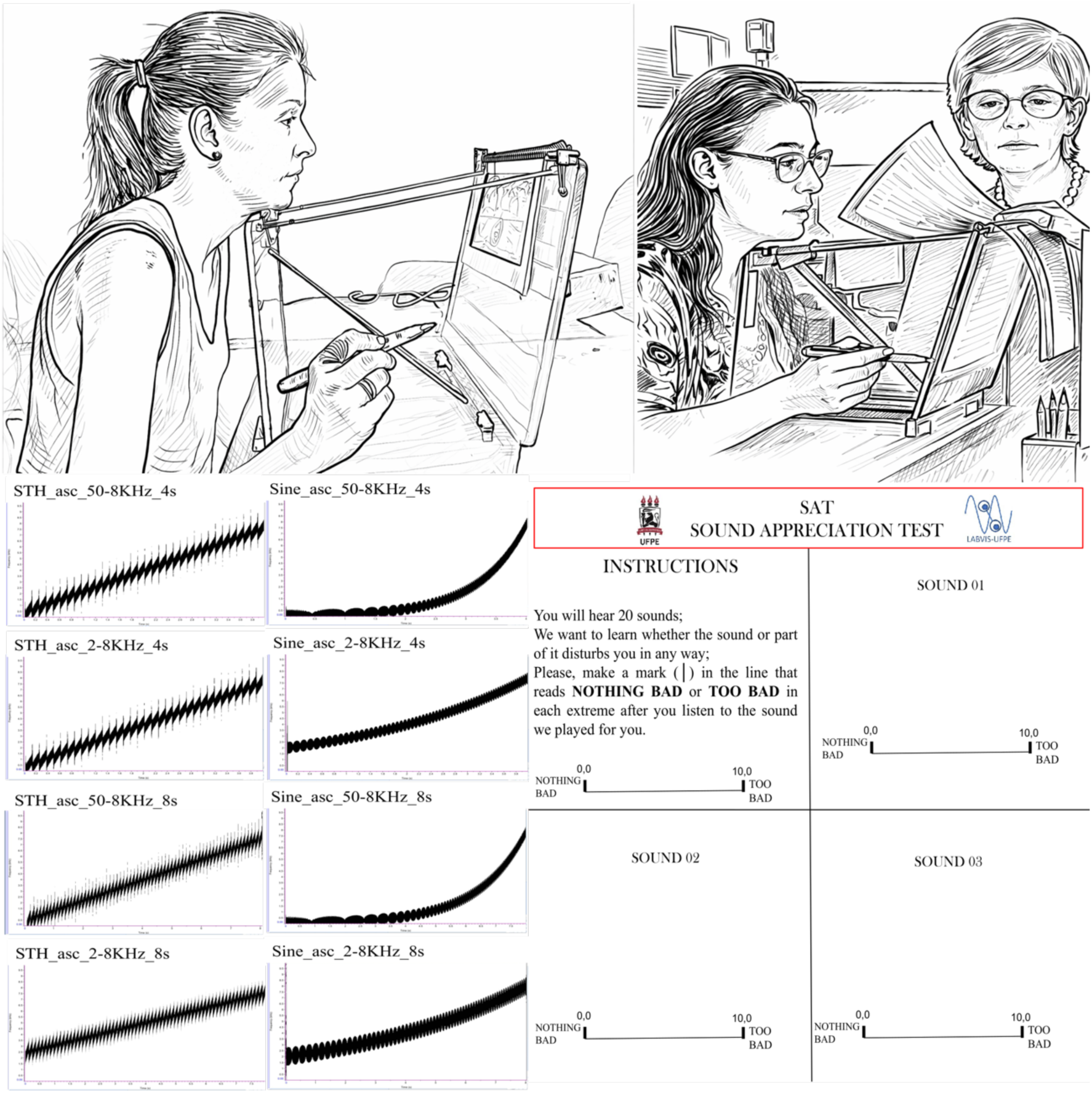
The Sound Appreciation Test (SAT) instruction page. Participants are asked to mark their level of auditory discomfort on a 10-cm visual analog scale (0 = NOTHING BAD, 10 = TOO BAD) after each of 20 sound stimuli (examples shown). The procedure includes a volume adjustment step to normalize individual auditory sensitivity.

#### (ii) The Sound Appreciation Test (SAT)

This version includes the same 16 pure-tone frequency sweeps already reported elsewhere^28–29^. Participants were asked to listen to each sound and mark on a pad the intensity of their perceived hearing discomfort. The folder contained written instructions and 20 pages, each featuring a 10 cm visual analog scale ranging from “NOTHING BAD” (left, no discomfort) to “TOO BAD” (right, maximum auditory discomfort). Each stimulus was played once through high-resolution headphones with noise attenuation, connected via Bluetooth to a cell phone at intensities of approximately 65 dB (half scale). Prior to measurements, to consistently normalize hearing sensitivities across participants, the volume was adjusted to each individual’s preferred listening level. This normalizes across individuals’ auditory sensitivity, so that discomfort ratings reflect each participant’s response intensity relative to their own baseline comfort level rather than absolute loudness.

### Procedure

After obtaining signed agreements from the public mental health institutions where the research took place and receiving approval from the Ethics Committee (Plataforma Brazil under CAEE registration numbers #23665419.5.0000.8807, #74757323.7.0000.5208, #57166622.0.0000.5208, #59516722.0.0000.5208, #70836723.0.0000.5208, and 70836723.0.3001.8807), individual experimental sessions began. The session sequence was identical for every experimental group and HC.

After a thorough explanation of the procedure and confirmation that the participant (and/or responsible sponsor) fully understood the research steps, written informed consent was obtained. This was followed by a sociodemographic questionnaire to record personal and family medical history, and by the Mini-Mental State Examination (MMSE). Whenever available, complementary data were obtained from medical records. Basic visual and auditory functions were then assessed before the PST and SAT measurements.

Instructions were printed and read aloud by the experimenter at the beginning of each psychophysical test. Each test started only after the experimenter was assured that the volunteer had fully understood the instructions. Further details can be found in the Supplementary material and in previous studies^28–29^.

### Statistical Analysis

Mann-Whitney U tests for independent samples were used to compare group characteristics and mean differences between experimental groups and their respective controls. Analyses of variance (ANOVA) were run individually for each patient group and their corresponding controls, with Bonferroni corrections for multiple comparisons. Normality tests were performed prior to parametric analyses. All statistical analyses were conducted using Statistica Ultimate Academic and GraphPad Prism 10.

## RESULTS

### Participant characteristics and group comparisons

Table 1 shows the relevant statistical profiles of participants. Mann-Whitney U tests for independent samples were used to assess group characteristics and mean differences between experimental groups and their respective controls. Sex distribution was unbalanced due to the predominantly male population of the participating mental health facilities. For differences in MMSE group scores, see Table 1 (and Supplementary Table 1).

Participants were matched for age with controls within each experimental group and within each sensory test. For SCHZ and BIP, all patients completed both PST and SAT; for FEP, the numbers differed slightly because not all patients performed both tests. Age means were exactly matched when rounded to integer. Formal education differed significantly between SCHZ and HC_SCHZ (p < 0.0001) and between BIP and HC_BIP (p < 0.05), but no such differences were found for FEP. Visual acuity was worse for SCHZ (p < 0.0001) and BIP (p < 0.001) relative to their controls, whereas FEP showed no difference. Smoking was strongly overrepresented in SCHZ compared to FEP and BIP (p < 0.0001). Regarding medication, only the active drug of antipsychotics were recorded.

### Raw data handling

Questionnaires and PST data were entered into Excel spreadsheets. PST values were the diameters of the circled areas (measured in centimeters) and converted to dva. Figure 2 illustrates raw data from one healthy control and one SCHZ participant. There were no missing data; differences in group sample sizes were treated by handling statistics of each experimental group separately against its own matched control group.

**Figure 2.**
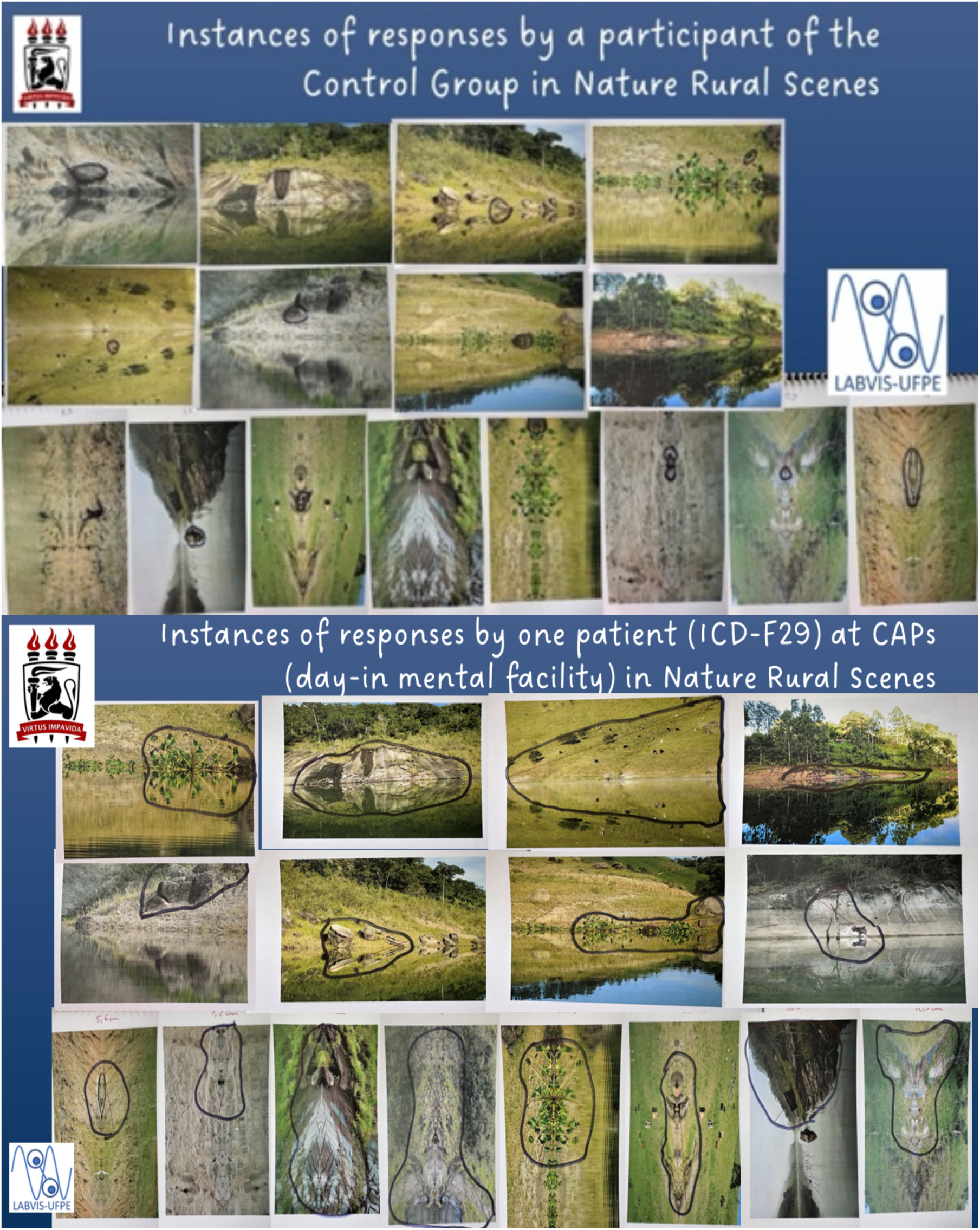
Representative PST responses. (Top) A healthy control participant circled a small area indicating normal pictorial size perception. (Bottom) A patient with first-episode psychosis (ICD-10 F29, unspecified psychosis) circled a substantially larger area, reflecting the enlarged visual size perception characteristic of schizophrenia-spectrum disorders. Circles were drawn by the participants in black ink.

SAT data are described as Sound Discomfort Level (SDL), rated on a continuous 0–10 visual analog scale. Sound stimuli were organized by modulation envelope (sawtooth, sine), order (ascending, descending), duration (4 s, 8 s), and frequency range (0.050–8 kHz, 2–8 kHz) for each participant and group.

### Main results

Group statistical analyses were carried out with Statistica Ultimate Academic and GraphPad Prism 10. Analyses of variance (ANOVA) were run individually for each patient group and respective controls, with Bonferroni corrections for multiple comparisons. Our main results are shown in Figures 3 and 4, organized as comparisons between each patient group and its age-matched HC.

**Figure 3.**
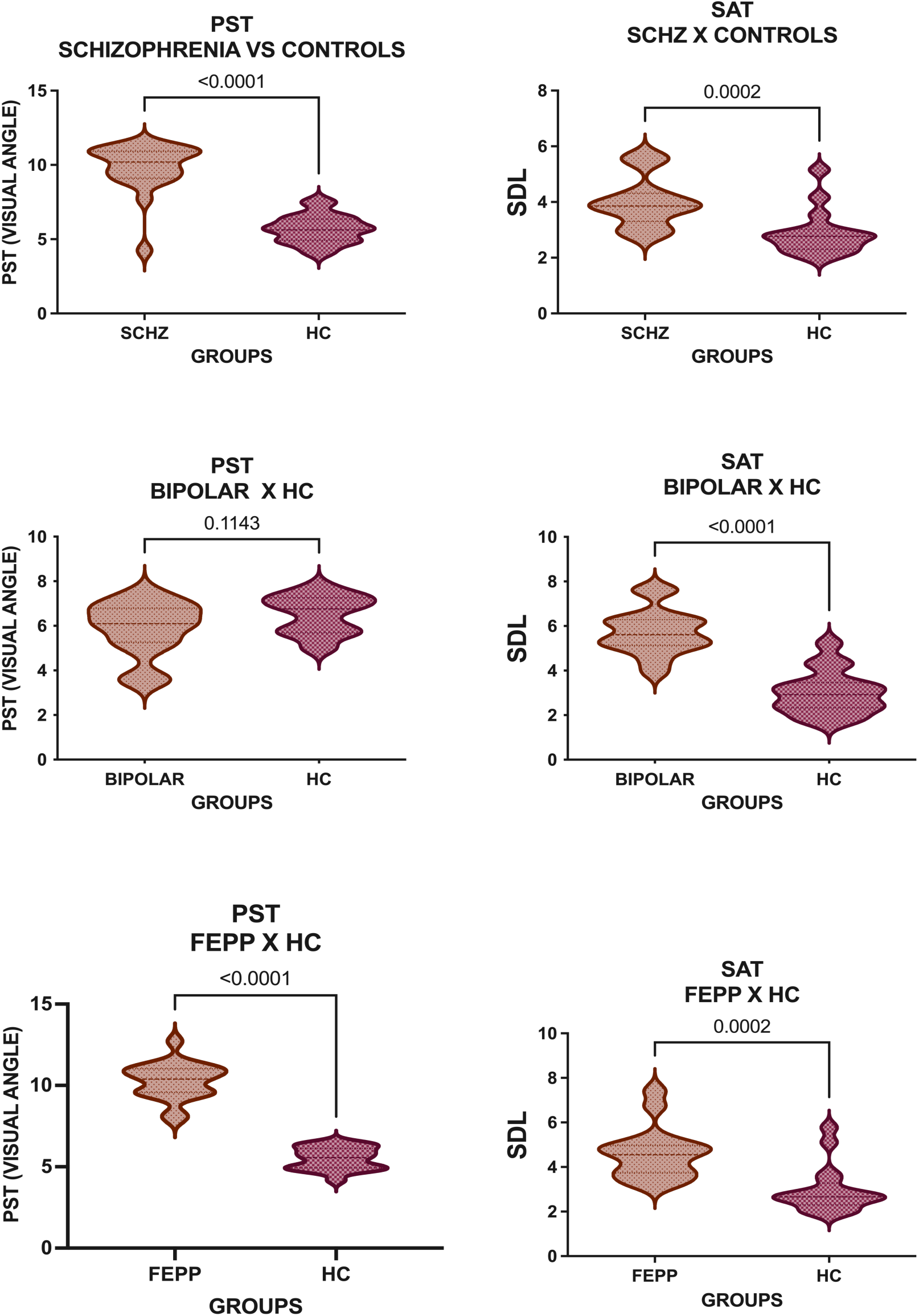
Group mean comparisons for PST (left column) and SAT (right column) across diagnostic groups. Error bars represent 95% confidence intervals. P-values from Mann-Whitney U tests (or t-tests) are shown. **Top row:** SCHZ vs. HC_SCHZ – PST p < 0.0001, SAT p = 0.0002. **Middle row:** BIP vs. HC_BIP – PST p = 0.1143 (non-significant), SAT p < 0.0001. **Bottom row:** FEP vs. HC_FEP – PST p < 0.0001, SAT p = 0.0002. Note the distinct pattern: SCHZ and FEP show enlarged PST responses, while BIP shows elevated SAT responses only.

**Figure 4.**
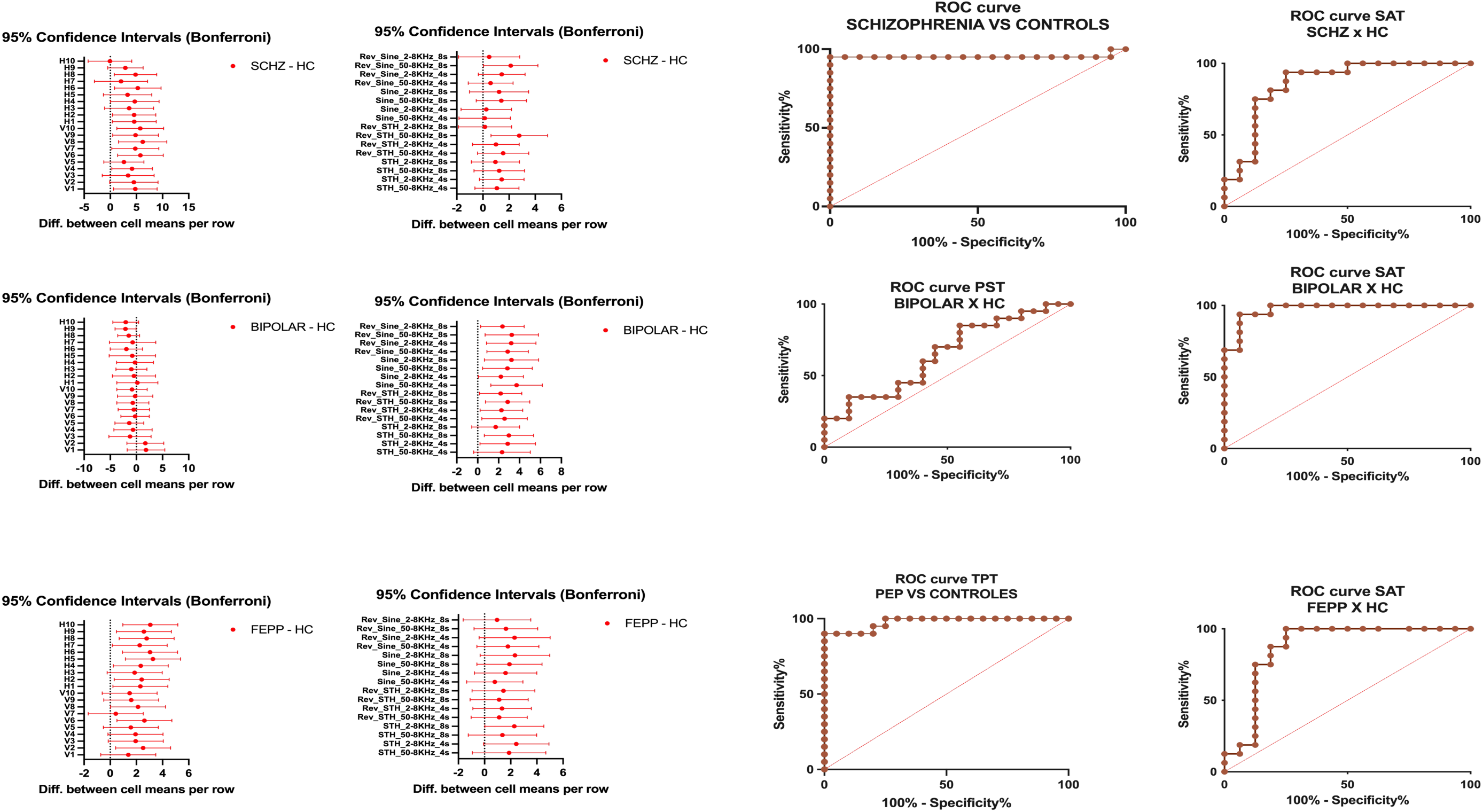
Left panels: 95% confidence intervals for PST (l) and SAT (middle left) after **Bonferroni correction** for multiple comparisons across the 20 stimuli (PST) or 16 sounds (SAT). The distribution of PST values for SCHZ and FEP is significantly above their respective controls, whereas BIP does not differ from HC_BIP. **Right panels:** Corresponding ROC curves for PST (middle right) and SAT (right) comparing each patient group with its age-matched control group. AUC values: PST – SCHZ: 0.953 (p < 0.0001), BIP: 0.648 (p = 0.1105), FEP: 0.978 (p < 0.0001); SAT – SCHZ: 0.863 (p = 0.0005), BIP: 0.973 (p < 0.0001), FEP: 0.871 (p = 0.0003). The diagonal dashed line represents chance (AUC = 0.500).

### Pictorial Size Test (PST)

Figure 3 (left) shows the first perceived pictorial size mean estimates and the statistical differences (Mann-Whitney U tests) for each experimental group versus its controls. SCHZ and FEP both circled significantly larger sizes than their respective HCs (p < 0.0001). BIP did not differ from HC_BIP.

Notably, the effect size for FEP (d = 2.86) was substantially larger than for chronic SCHZ (d = 0.63), suggesting that the visual size alteration may be most pronounced in the early stages of schizophrenia-spectrum disorders regardless of normal visual acuity.

### Sound Appreciation Test (SAT)

Figure 3 (right) shows mean SDL. All three patient groups reported significantly higher auditory discomfort than their respective controls: BIP (p < 0.0001), FEP (p < 0.0002), and SCHZ (p < 0.0002). Supplementary Figure 1 provides group means for each measurement and shows each significant difference found for specific sound sweeps.

### Simultaneous parametric statistical treatments

To examine possible image symmetry effects in PST, three-factor ANOVAs (group × symmetry × scene) were performed separately for SCHZ, BIP and FEP after normality testing.

Significant group differences were found for SCHZ vs HC_SCHZ (F₁,₄₄ = 13.37, p < 0.0007) and for FEP vs HC_FEP (F₁,₃₈ = 12.81, p < 0.0010), but not for BIP. A light symmetry effect was present in the BIP group (F₁,₃₅ = 4.629, p < 0.0384). No symmetry × group interactions were observed for SCHZ or FEP; however, a three-way interaction (group × symmetry × scene) was found for SCHZ (F₉,₃₉₆ = 3.236, p = 0.0008). Supplementary Figure 2 illustrates differences in first pictorial sizes and symmetries for SCHZ and BIP.

For SAT, two-factor ANOVAs revealed significant group differences for all three comparisons: SCHZ vs HC_SCHZ (F₁,₄₄ = 7.619, p < 0.0084), FEP vs HC_FEP (F₁,₃₇ = 7.609, p < 0.0090), and BIP vs HC_BIP (F₁,₃₅ = 30.39, p < 0.0001), with the largest magnitude for BIP. A Group × Sound interaction was found only for SCHZ (F₁₅,₆₆₀ = 2.101, p < 0.0086).

Supplementary Figure 1 (bottom panels) shows that most PST values for SCHZ and FEP lie well above their respective HC 95 % confidence intervals, whereas BIP differs from HC_BIP in only 5 out of 20 stimuli.

### Bonferroni corrections

Figure 4 (left and middle left) displays 95 % confidence intervals with Bonferroni corrections for PST (left) and SAT (middle left). The distribution of first perceived pictorial sizes in SCHZ and FEP remains very similar, but the magnitude is higher for FEP. Values for BIP do not deviate from zero.

### Receiver Operating Characteristic and power analyses

Figure 4 (right panels) shows ROC curves for each patient group versus respective controls. PST distinguished SCHZ (AUC = 95.3 %, p < 0.0001) and FEP (AUC = 97.8 %, p < 0.0001) from their controls, but not BIP (AUC = 64.8 %, p = 0.1105). SAT distinguished BIP extremely well (AUC = 97.3 %, p < 0.0001), but distinguished SCHZ (AUC = 86.3 %, p < 0.0005) and FEP (AUC = 87.1 %, p < 0.0003) from controls less efficiently than PST for both groups.

Power analysis (refer to Supplementary Table 2) showed that PST has 95 % power (d = 0.63, p < 0.0001) to distinguish SCHZ from HC, and 99 % power (d = 2.86, p < 0.0001) to distinguish FEP from HC. SAT has 99 % power (d = 2.73, p < 0.0001) to distinguish BIP from HC, and slightly lower power for SCHZ (99 %, d = 1.29, p < 0.0005) and FEP (99 %, d = 1.46, p < 0.0003).

### Development of a sensory-based classification algorithm (SoundPics)

Given the strong statistical effects and the coupled specificity of PST and SAT, we developed a hierarchical rule-based algorithm (SoundPics) derived exclusively from the SCHZ and BIP chronic patients data. The algorithm uses eight hierarchical rules with empirically defined cut-offs based on observed patient thresholds (Supplementary Table 3).

Compared to an earlier version, SoundPics V3 improved overall accuracy from 65.7% to 70.8% and bipolar sensitivity from 30.0% to 46.9% (Supplementary Table 4). The algorithm returned ‘undetermined’ for 11.1% of cases (9 of 81), indicating that sensory profiles were inconclusive for differential diagnosis in a small subset of patients.

SoundPics V3 achieved 70.8% accuracy among clinically determined cases (72 of 81 cases), compared with 66.7% for linear discriminant analysis (LDA) applied to the same data (Supplementary Table 5). The ROC curve for the coupled SoundPics algorithm yielded an AUC of 0.735, approaching the performance of LDA (AUC 0.760; Supplementary Figure 5).

In an independent sample of 17 FEP patients (not part of the main FEP group), the SoundPics algorithm showed a sensitivity of 90% for SCHZ and 69.2% agreement regarding provisional diagnoses. When we included in the sample those who participated in the present study (N=39 + N=17, total N=56), the overall agreement with established clinical diagnoses was 64.7%, and for SCHZ alone it was 76.8%. Further details are found in the Supplementary material in the session “Differential classification between SCHZ, BIP and undetermined”.

### External validation using the FEP group as an independent test set

The psychophysical results from the FEP group provide convergent validity for the SCHZ and BIP group findings. ROC profiles for both PST and SAT were very similar between FEP and SCHZ groups, even though measurements were totally independent. These are not predictions from latent constructs or psychometric resources, but direct sensory psychophysical measurements from FEP patients that corroborate the SCHZ and BIP group findings. Notably, the FEP sample was recruited from AMPEP-EBSERH-HC-UFPE, an outpatient service that selects patients clinically pre-classified as possibly developing schizophrenia.

## Discussion

This study introduces a novel psychophysical approach to early psychosis evaluation, focusing on sensory processing rather than cognition or behavior alone. The present findings should be interpreted within a sensory psychophysics framework, in which sensory responses are directly measured rather than inferred from latent variables. This distinction is critical, as it positions PST and SAT as objective sensory probes that access early-stage processing alterations, potentially preceding and complementing traditional clinical assessments. Our main findings are: (1) patients with schizophrenia (SCHZ) and first-episode psychosis (FEP) show augmented visual size perception on the Pictorial Size Test (PST), while bipolar disorder (BIP) patients do not; (2) all three patient groups report elevated auditory discomfort on the Sound Appreciation Test (SAT), with the largest effect in BIP; and (3) a hierarchical rule-based algorithm (SoundPics) coupling PST and SAT results can differentiate SCHZ from BIP with 70.8% accuracy among clinically determined cases, leaving 11.1% as undetermined.

These results demonstrate that low-cost psychophysical measurements can reveal distinct sensory profiles in early psychosis that are not captured by standard clinical interviews. Below, we discuss the implications of these findings, their alignment with current literature, and directions for future research.

### Visual size alterations in schizophrenia and first-episode psychosis

Our finding of strong PST alterations in SCHZ is consistent with recent evidence showing anatomical and functional differences in the retina of people with schizophrenia^32,40–44^. The augmented perceived size (measured in degrees of visual angle) may reflect early sensory processing deficits, possibly involving a shift of the contrast sensitivity function to lower spatial frequencies, inhibitory effects affecting surround retinal suppression, and alterations of cortical magnification mechanisms.

Importantly, the effect size was substantially larger in FEP (d = 2.86) than in chronic SCHZ (d = 0.63) despite normal acuity found in the FEP group. In SoundPics, the PST effect appears independent of visual acuity since FEP patients had normal acuity and showed the largest effect, while SCHZ patients had worse acuity but showed smaller effects. This dissociation argues against a simple sensory degradation account and supports a central perceptual alteration not related to age but to the disease itself.

This suggests that the visual size alteration may be most pronounced in the early stages of schizophrenia-spectrum disorders, potentially attenuating with illness progression or chronic antipsychotic treatment. Longitudinal studies are in progress to test this hypothesis.

### Auditory discomfort across diagnostic groups

All three patient groups reported significantly higher auditory discomfort than healthy controls on the SAT, but the pattern differed: BIP showed the largest effect (d = 2.73), while SCHZ and FEP had smaller but still significant effects (d = 1.29 and 1.46, respectively). This finding aligns with clinical observations that individuals with bipolar disorder often report heightened sensitivity to sounds during mood episodes, but our data suggest that elevated auditory discomfort may be a trait-like feature present even in clinically stable chronic patients.

The SAT procedure – adjusting volume to each participant’s preferred listening comfort level before stimulus presentation – normalizes for differences in individuals’ auditory sensitivities. Thus, the discomfort ratings reflect perceptual responses to the frequency sweeps relative to each person’s own baseline, not absolute loudness. This methodological choice strengthens the validity of cross-group comparisons.

### Distinct sensory profiles for schizophrenia and bipolar disorder

The dissociable patterns observed here (visual alteration predominant in SCHZ/FEP; auditory discomfort predominant in BIP) suggest that sensory testing may assist in early differential diagnosis. While both PST and SAT detect alterations in schizophrenia, PST shows stronger effects for SCHZ, and SAT shows stronger specificity for BIP (Figures 3 and 4; Supplementary Figures).

These profiles are not merely artifacts of medication or chronicity. All SCHZ and BIP patients were medicated, yet the patterns remained distinct. Moreover, the FEP group – all of whom met ICD-11 criteria for first-episode psychosis and were clinically pre-classified as suspected schizophrenia at the time of testing, being recruited from AMPEP (an ambulatory dedicated to presumed schizophrenia) – showed a visual profile remarkably similar to chronic SCHZ. This argues against a primary medication effect and supports the utility of PST as an early indicator of schizophrenia-spectrum risk.

### The SoundPics algorithm: towards clinical translation

Given the coupled specificity of PST and SAT, we developed a hierarchical rule-based algorithm (SoundPics) derived exclusively from the chronic SCHZ and BIP data. All chronic SCHZ and BIP patients had established diagnoses according to ICD-11 criteria (schizophrenia and bipolar disorder, respectively). Compared to an earlier version, SoundPics V3 improved overall accuracy from 65.7% to 70.8% and bipolar sensitivity from 30.0% to 46.9% (Supplementary Table 4). The algorithm returns “undetermined” for 11.1% of cases – an honest reflection of its limitations, which may be acceptable in clinical settings where ambiguous cases require further assessment.

SoundPics V3 achieved 70.8% accuracy among clinically determined cases (72 of 81), compared with 66.7% for linear discriminant analysis (LDA) applied to the same data (Supplementary Table 5). The ROC curve for the coupled algorithm yielded an AUC of 0.735, approaching LDA (AUC 0.760; Supplementary Figure 5). While not yet ready for standalone use, this proof-of-concept demonstrates that sensory psychophysics can contribute to diagnostic decision-making.

### External validity and the FEP sample

A particularly important aspect of the present findings is that the FEP group functioned as an independent external validation sample. Despite being recruited from a distinct clinical setting and evaluated independently from chronic patients, FEP participants exhibited sensory profiles closely matching those observed in schizophrenia. This convergence strengthens the robustness of the findings and suggests that the observed visual alterations are not restricted to chronic stages but may emerge early in the development of the disorder.

In this study the FEP group was recruited from AMPEP-EBSERH-HC-UFPE, an outpatient service that specifically selects patients with a first episode of psychosis and a clinical suspicion of schizophrenia (rather than bipolar disorder). All FEP patients met ICD-11 criteria for first-episode psychosis and were being followed with the working hypothesis that they would likely develop schizophrenia. So, the FEP sample was independent from the chronic SCHZ group (which came predominantly from a forensic hospital and CAPS units). The fact that FEP patients showed PST and SAT profiles nearly identical to those of chronic SCHZ – despite lacking a final diagnosis of schizophrenia at the time of testing – provides strong convergent and external validity for the algorithm’s schizophrenia-spectrum classification. However, additional external validation in an entirely independent cohort from a different geographical region should reinforce this findings before clinical implementation.

### Clinical implications

The following practical implications can be drawn for clinicians and researchers:

- **Screening tool:** The SoundPics battery (PST + SAT) takes approximately 15 minutes to administer and requires only a printed folder, headphones, and a cell phone. It could be used in primary care or community mental health settings to identify individuals with possible early psychosis who warrant specialized assessment.
- **Diagnostic adjunct:** For patients already under evaluation, SoundPics provides adjunctive information about sensory processing not captured by standard scales (PANSS, YMRS, etc.). A PST score greater than specific numbers of degrees of visual angle combined with given values of SAT scores (exact cut-offs to be refined in future studies) suggests a schizophrenia-spectrum profile; a SAT score greater than specific values coupled with low PST values suggests a bipolar profile. These are not substitutes for clinical diagnosis (which remains based on ICD-11/DSM-5 criteria) but can further clinical judgement.
- **Risk stratification:** The finding that FEP patients with suspected schizophrenia show larger visual effects than chronic SCHZ raises the possibility that PST could help identify individuals at ultra-high risk who might benefit from early intervention prior to cognitive deficits. Longitudinal follow-up of the FEP cohort is ongoing to test this hypothesis.
- **Monitoring treatment response:** Although preliminary, the apparent stability of sensory profiles across medicated and unmedicated patients suggests that these

measures may be trait-like. If confirmed, they could serve as stable markers for diagnostic classification rather than state markers of symptom fluctuation.

### Study limitations

We acknowledge several limitations:

1. **Geographic and sample restrictions:** All participants were recruited from public mental health facilities in Pernambuco, Brazil. Findings may not generalize to other populations or healthcare systems.
2. **Inclusion of a forensic subsample:** Some SCHZ patients came from a forensic psychiatric hospital (HCTP), which may differ from community-dwelling patients in illness severity or other factors.
3. **Sex imbalance:** Sex distribution was unbalanced (predominantly male), reflecting the characteristics of the participating facilities. We could not analyze sex-specific effects.
4. **No standardized symptom scales:** While we administered the MMSE for cognitive screening, we did not collect conventional symptom severity measures (PANSS, YMRS, BPRS). This limits our ability to correlate perceptual alterations with clinical states such as positive symptoms or mood episode phases.
5. **Medication effects not systematically assessed:** Although our data suggest that medication does not strongly interfere with the results (consistent patterns across medicated and partially medicated groups), we did not correlate dosages, durations, or specific antipsychotic types with SoundPics data. Ongoing studies are addressing this.
6. **Lack of test-retest reliability data:** We have not yet assessed the variability of PST and SAT scores with medication dosage over time within individuals. We have a study in progress evaluating this.
7. **FEP selection bias:** The FEP group was recruited from an ambulatory that screens and treats patients with suspected schizophrenia. Therefore, our findings cannot be generalized to all first-episode psychosis patients (e.g., those with suspected bipolar disorder). This also means that FEP patients from another ambulatory that screens for bipolar disorder should serve as complementary external validity. Ongoing studies include FEP patients from both suspected schizophrenia and suspected bipolar disorder ambulatories.

On the other hand, some features that could be seen as limitations also represent strengths: the naturalistic public health setting enhances ecological validity; the inclusion of multiple sites and different time periods (2019-2025) supports robustness; and the consistent results across diverse patient subgroups argue for generalizability within the Brazilian public mental health system.

### Future directions

We have ongoing studies at the twin ambulatories (AMPEP for presumed schizophrenia, AMBIP for presumed bipolar disorder) specifically investigating: (i) the relationship between sensory alterations and medication type/dosage; (ii) the concordance between psychophysical measurements and the attending clinician’s impression at the time of testing; and (iii) a three-year follow-up to improve the SoundPics algorithm and test its predictive value for diagnostic stability.

We should note that evaluating the SoundPics battery (PST and SAT) with AMBIP patients – i.e., first-episode psychosis patients with a clinical suspicion of bipolar disorder – to further test our hypothesis that visual size perception is not as strongly affected in bipolar-spectrum early psychosis as it is in SCHZ early psychosis, and that auditory discomfort remains elevated regardless of medication and disease duration, would provide strong evidence for the differential diagnostic utility of SoundPics from the first episode onward.

Our ultimate objective is to be able to medicate sensory symptoms even prior to cognitive dysfunction, and we are submitting a project to the ethics committee to investigate this possibility.

## CONCLUSION

The present study introduces low-cost psychophysical measurements (PST and SAT) that reveal distinct sensory profiles in early psychosis. Schizophrenia and first-episode psychosis with suspected schizophrenia are characterized by augmented visual pictorial size perception, while bipolar disorder is characterized by pronounced auditory discomfort. The hierarchical SoundPics algorithm, coupling both tests, achieved promising accuracy (70.8% among clinically determined cases) for differentiating schizophrenia from bipolar disorder. Importantly, the replication of schizophrenia-like sensory profiles in an independent FEP sample supports the external validity of the proposed framework and highlights its potential relevance for early-stage investigation of psychosis. By capturing perceptual alterations not readily apparent in standard clinical interactions, this approach may ultimately contribute to earlier and more accurate differential diagnosis. Together, these findings suggest that sensory testing can serve as a screening tool and provide diagnostic adjunctive information in early psychosis, particularly when clinical presentation is ambiguous and taken together with other clinical tools^45–47^.

## Data Availability

We are attaching all supplementary data including raw data files.
Sound files are found in: fonae.org/audios

## ACKNOWLEDGEMENTS

We thank the Coordenação de Aperfeiçoamento de Pessoal do Nível Superior (CAPES) for support from the PROAP program and for the graduate fellowships to Mateus Monteiro de Gois Barros, Katherine Karla Souza da Silva, and Amanda Almeida Rodrigues e Silva, and PROPESQI-UFPE for the Scientific Initiation scholarship to Kayke Kauã de Siqueira. We also thank Maria Cristina Fortes Santos de Bustamante for making freely available a collection of photographs from her own authorship. We particularly thank Professor José Mauricio Haas Bueno for his comments and review of the manuscript.

## DISCLOSURE

None of the authors have conflict of interest.

## AUTHOR CONTRIBUTIONS

Maria Lucia de Bustamante Simas *Idealized instruments, designed experiments, wrote the manuscript and performed statistics treatment and illustrations of data*. *All data are available under request*, raw data are provided with Supplemental material.

Aline Mendes Lacerda *supervised part of data collection*

Joselma Tavares Frutuoso *supervised part of data collection*

Isabela de Fatima Pina de Almeida *Head of AMPEP public mental health service; supervisor & Clinical assessments*

Mateus Monteiro de Gois Barros *provided collected data from his master’s thesis, prepared references, helped preparing documents for the ethics committee*.

Katherine Karla Souza da Silva *provided collected data from her master’s thesis*.

Thays Macambira da Silva *provided collected data from her master’s thesis*.

Giuliano Baltar Melo de Souza Ramos *provided collected data from his master’s thesis*

Tamires Lima da Silva *provided collected data from her master’s thesis*

Naianna Mocelin Ribeiro dos Santos *provided collected data from her master’s thesis*.

Amanda Almeida Rodrigues e Silva *collected data, prepared references*.

Kayke Kauã de Siqueira *collected data*.

## Notes

### Competing Interest Statement

The authors have declared no competing interest.

### Funding Statement

We only received funds from the Coordenacao de Aperfeicoamento de Pessoal de Nivel Superior through the graduate research program for printing TEST material

### Author Declarations

All experimente were approved by the Ethics Committee from the Federal University of Pernambuco and University Hospital das Clinicas and Plataforma Brazil under CAEE registration numbers 23665419.5.0000.8807, 74757323.7.0000.5208, 57166622.0.0000.5208, 59516722.0.0000.5208, 70836723.0.0000.5208, 70836723.0.3001.8807

### Summary of Updates

This version was reformulated to better convey information about the same set of experiments focusing on the possible use of SoundPics for psychiatric and pschological screening together with present scales on use.

## REFERENCES

1. Akudjedu TN, Tronchin G, McInerney S, Scanlon C, Kenney JP, McFarland J, et al. Progression of neuroanatomical abnormalities after first-episode of psychosis: A 3-year longitudinal sMRI study. J Psychiatr Res. 2020;130:137–51.

2. Briend F, Nelson EA, Maximo O, Armstrong WP, Kraguljac NV, Lahti AC. Hippocampal glutamate and hippocampus subfield volumes in antipsychotic-naive first episode psychosis subjects and relationships to duration of untreated psychosis. Transl Psychiatry. 2020;10:137.

3. Garcia-Marti G, Escarti MJ, Nacher J, Perez-Rando M, Mane A, Usall J, et al. Progressive loss of cortical gray matter in first episode psychosis patients with auditory hallucinations. Schizophr Res. 2024;267:534–45.

4. Chopra S, Levi PT, Holmes A, Orchard ER, Segal A, Francey SM, et al. Brainwide Anatomical Connectivity and Prediction of Longitudinal Outcomes in Antipsychotic-Naïve First-Episode Psychosis. Biol Psychiatry. 2024.

5. Blasco MB, Aji KN, Ramos-Jiménez C, Leppert IR, Tardif CL, Cohen J, et al. Synaptic Density in Early Stages of Psychosis and Clinical High Risk. JAMA Psychiatry. 2024.

6. Berthet P, Haatveit BC, Kjelkenes R, Worker A, Kia SM, Wolfers T, et al. A 10-Year Longitudinal Study of Brain Cortical Thickness in People with First-Episode Psychosis Using Normative Models. Schizophr Bull. 2024;sbae107.

7. Bernstein HG, Nussbaumer M, Vasilevska V, Dobrowolny H, Nickl-Jockschat T, Guest PC, et al. Glial cell deficits are a key feature of schizophrenia: implications for neuronal circuit maintenance and histological differentiation from classical neurodegeneration. Mol Psychiatry. 2024.

8. Gao Z, Xiao Y, Zhu F, Tao B, Zhao Q, Yu W, et al. Multilayer network analysis reveals instability of brain dynamics in untreated first-episode schizophrenia. Cereb Cortex. 2024;34:bhae402.

9. Yang C, Zhang W, Yao L, Liu N, Shah C, Zeng J, et al. Functional Alterations of White Matter in Chronic Never-Treated and Treated Schizophrenia Patients. J Magn Reson Imaging. 2017;46:757–64.

10. Anteraper SA, Guell X, Collin G, Qi Z, Ren J, Nair A, et al. Abnormal Function in Dentate Nuclei Precedes the Onset of Psychosis: A Resting-State fMRI Study in High-Risk Individuals. Schizophr Bull. 2021;47:1421–30.

11. Ruan X, Zhang L, Duan M, Yao D, Luo C, He H. Disrupted functional connectivity between visual and emotional networks in psychosis risk syndromes through representational similarity analysis. Front Psychiatry. 2025;16:1533675.

12. Rodríguez-Toscano E, Martínez K, Fraguas D, Janssen J, Pina-Camacho L, Arias B, et al. Prefrontal abnormalities, executive dysfunction and symptoms severity are modulated by COMT Val158Met polymorphism in first episode psychosis. Rev Psiquiatr Salud Ment (Barc). 2022;15:74–87.

13. Howes OD, Bukala BR, Beck K. Schizophrenia: from neurochemistry to circuits, symptoms and treatments. Nat Rev Neurol. 2024.

14. Squarcina L, Stanley JA, Bellani M, Altamura CA, Brambilla P. A review of altered biochemistry in the anterior cingulate cortex of first-episode psychosis. Epidemiol Psychiatr Sci. 2017;26:122–8.

15. Berdeville CHSF, Silva-Amaral D, Dalgalarrondo P, Banzato CEM, Martins-de-Souza D. A scoping review of protein biomarkers for schizophrenia: State of progress, underlying biology, and methodological considerations. Neurosci Biobehav Rev. 2025;168:105949.

16. American Psychiatric Association. Diagnostic and statistical manual of mental disorders. 5th ed., text revision. American Psychiatric Association. 2022.

17. World Health Organization. International classification of diseases for mortality and morbidity statistics. 11th ed. World Health Organization. 2022.

18. Tandon R, Nasrallah H, Akbarian S, Carpenter WT Jr, DeLisi LE, Gaebel W, et al. The schizophrenia syndrome, circa 2024: What we know and how that informs its nature. Schizophr Res. 2024;264:1–28.

19. Lima MS, Soares BG, Paoliello G, Vieira RM, Martins CM, Mota Neto JI, et al. The Portuguese version of the Clinical Global Impression – Schizophrenia Scale: validation study. Rev Bras Psiquiatr. 2007;29:246–9.

20. Overall JE, Gorham DR. The brief psychiatric rating scale. Psychol Rep. 1962;10:799–812.

21. Kay SR, Fiszbein A, Opler LA. The Positive and Negative Syndrome Scale (PANSS) for Schizophrenia. Schizophr Bull. 1987;13:261–76.

22. Yung AR, Yuen HP, McGorry PD, Phillips LJ, Kelly D, Dell’Olio M, et al. Mapping the onset of psychosis: the Comprehensive Assessment of At-Risk Mental States. Aust N Z J Psychiatry. 2005;39:964–71.

23. Carvalho VA, Caramelli P. Brazilian adaptation of the Addenbrooke’s Cognitive Examination-Revised (ACE-R). Dement Neuropsychol. 2007;1:212–6.

24. Folstein MF, Folstein SE, McHugh PR. “Mini-mental state”. A practical method for grading the cognitive state of patients for the clinician. J Psychiatr Res. 1975;12:189–98.

25. Nasreddine ZS, Phillips NA, Bédirian V, Charbonneau S, Whitehead V, Collin I, et al. The Montreal Cognitive Assessment, MoCA: a brief screening tool for mild cognitive impairment. J Am Geriatr Soc. 2005;53:695–9.

26. Simas MLDB, Nogueira RMTBL, Menezes GMM, Amaral VF, Lacerda AM, Santos NA. O uso de pinturas de Dalí como ferramenta para avaliação das alterações na percepção de forma e tamanho em pacientes esquizofrênicos. Psicol USP. 2011;22:67–80.

27. Lacerda, AM, Simas, MLB, Menezes, GMM. (2020). Changes in visual size perception in schizophrenia and depression. Psicol Pesquisa, 14 (spe), 140–153.

28. Simas MLDB, Maranhão ACT, Lacerda AM, Teixeira FS, Freire CHR, Raposo CCS, et al. Pictorial size perception in schizophrenia. Psicol Reflex Crit. 2021;34:36.

29. Simas MLDB, Santos NRM, Lacerda AM. Auditory perceptual discomfort and low-hearing tolerance in the first episode psychosis. Psicol Reflex Crit. 2022;35:20.

30. Simas MLDB, Silva TL, Santos NRM, Lacerda AM. Mutually exclusive disorder-dependent hearing discomfort in first-episode psychosis and panic disorder: two experiments using the same auditory stimulus set and two similar musical sequences. Psicol Reflex Crit. 2022;35:37.

31. Haigh SM, Haggerty JA, Delgado A. Auditory discomfort and visual sensitivity. Vision Res. 2025;234:108655.

32. Ifrah C, Herrera SN, Silverstein SM, Corcoran CM, Gordon J, Butler PD, et al. The Relationship between Clinical and Psychophysical Assessments of Visual Perceptual Disturbances in Individuals at Clinical High Risk for Psychosis: A Preliminary Study. Brain Sci. 2024;14:819.

33. Simas MLDB, Esquizofrenia: seus fenômenos perceptivos e cognitivos na primeira pessoa. Editora Appris, 2018.

34. Santos NRM. Avaliação de alterações sensório-perceptuais de pacientes em primeiro episódio psicótico. [Dissertação de mestrado]. Recife: Universidade Federal de Pernambuco; 2021.

35. Silva TL. Desconforto sonoro em pessoas com transtorno de pânico e autorrelato de sintomas sensoriais concomitantes. [Dissertação de mestrado]. Recife: Universidade Federal de Pernambuco; 2022.

36. Ramos GBMS. Avaliação multissensorial de usuários de Centro de Atenção Psicossocial com foco nos diagnósticos que incluem sintomas de psicose. [Dissertação de mestrado]. Recife: Universidade Federal de Pernambuco; 2023.

37. Barros MMG. Percepção pictorial, força de preensão palmar e nível de desconforto sonoro na esquizofrenia. [Dissertação de mestrado]. Recife: Universidade Federal de Pernambuco; 2023.

38. Silva TM. Aspectos sensório-perceptuais nas condições pós-COVID. [Dissertação de mestrado]. Recife: Universidade Federal de Pernambuco; 2024.

39. Silva KKS. Avaliação multissensorial em pessoas com transtorno bipolar. [Dissertação de mestrado]. Recife: Universidade Federal de Pernambuco; 2024.

40. Kapisthalam S, Bi H, Zhang Y, Clark AM, Thompson JL, Poletti M, et al. Fixational eye movements and visual acuity in patients with schizophrenia. J Vis. 2025;25:2442.

41. Shoham N, Lewis G, Hayes JF, Silverstein SM, Cooper C. Association between visual impairment and psychosis: A longitudinal study and nested case-control study of adults. Schizophr Res. 2023;254:81–89. doi:10.1016/j.schres.2023.02.017.

42. Green KM, Choi JJ, Ramchandran RS and Silverstein SM (2022) OCT and OCT Angiography Offer New Insights and Opportunities in Schizophrenia Research and Treatment. Front. Digit. Health 4:836851. doi: 10.3389/fdgth.2022.836851

43. Schwarzer JM, Meyhoefer I, Antonucci LA, et al. The impact of visual dysfunctions in recent-onset psychosis and clinical high-risk state for psychosis. Neuropsychopharmacology. 2022;47(12):2051–2060. doi:10.1038/s41386-022-01385-3

44. Diamond, A., Silverstein, S.M. & Keane, B.P. Visual system assessment for predicting a transition to psychosis. Transl Psychiatry 12, 351 (2022). 10.1038/s41398-022-02111-9

45. Williams TF, Gold JM, Waltz JA, Schiffman J, Ellman LM, Strauss GP, et al. Identifying individuals at clinical high risk for psychosis using a battery of tasks sensitive to symptom mechanisms. Transl Psychiatry. 2025;15:311.

46. Gadelha A, Biokino RM, Lorencetti P, Crossley NA, Bordini D, Massuda R. Introducing a new severity specifier for schizophrenia: conceptual framework and clinical implications. Braz J Psychiatry. 2024;46:e20243722.

47. Jobim GS, do Amaral JV, Pacheco JPG, Gadelha A, Miguel EC, Bressan RA, et al. Clinical properties of the short mood and feelings questionnaire: Development of a free calculator based on the Brazilian high-risk cohort study. J Psychiatr Res. 2025;190:457–464.

