## Supplementary material for "SoundPics: visual and auditory psychophysics reveals distinct sensory profiles that differentiate schizophrenia from bipolar disorder in early psychosis across three independent Brazilian public health clinical samples": suplementary material

***Title:***

### 1. Complementary information to METHOD

#### 1.1 Supplementary Table 1

Table 1 shows scores on neuropsychological tests from experimental and healthy control groups. MMSE was available for 29 of 36 BIP patients and 29 of 36 HC-BIP; full sample sizes reported in main Table 1.

| GROUP | N | MMSE Median | *p* VALUE | N | ADDENBROOKE (ACE-R) | *p* VALUE |
|---|---|---|---|---|---|---|
| **FEP / HC-FEP** | 39 | 30 / 30 | NS | 14 | 80.5 / 91.4 | 0.0155 |
| **SCHZ / HC-SCHZ** | 42 | 21 / 30 | < 0.0001 | — | — | — |
| **BIPOLAR / HC-BIP** | 29 | 23 / 29 | < 0.0001 | — | — | — |
| **SCHZ / BIP** | 42 / 29 | 21 / 23 | 0.0069 | — | — | — |

***Note.*** MMSE = Mini-Mental State Examination; ACE-R = Addenbrooke's Cognitive Examination; FEP = first-episode psychosis; SCHZ = schizophrenia; BIP = bipolar disorder; HC = healthy controls; NS = not significant.

#### 1.2 Screening procedures

***Other Equipment and Instruments***

The experiment also involved the following equipment and instruments:

a. **Background questionnaire** – Sociodemographic data and medical background from patients were obtained either from medical records (with consent) and/or structured questionnaires.

b. **Screening test for cognition** – Cognitive functioning was evaluated with the Brazilian adaptation of the Mini Mental State Examination (MMSE) (Santos, 2010).

c. **Screening test for visual acuity** – The Android/iOS app *Smart Optometry* (by Smart Optometry) with varying sizes of Snellen "E" stimuli were presented either on cellphones or 8" tablets. Participants were asked to indicate the open side of the "E" either orally or by hand gesture. When more than 50% errors were made within the same size, the acuity corresponding to the size immediately prior to that was considered.

d. **Screening test for hearing** – Either the application *Test of audition* (developed by e-audiologia for Android) or *EasyHearingTest* (developed by Hiroaki Ito for iOS) was used to evaluate bilateral hearing of participants. A few participants wearing hearing aids and presenting expected age-standard hearing results were included in the study.

### 2. Complementary information to RESULTS

#### 2.1 Supplementary Table 2. PST and SAT statistical power, Cohen's d, and 95% CI with respective observed probabilities for SCHZ, BIP and FEP.

PST = Pictorial Size Test score (degrees of visual angle, *dva*). SAT = Sound Appreciation Test score (0–10 sound discomfort level scale – arbitrary units).

| GROUP | Test | AUC | *d* | Power | CI 95% | *p*-value |
|---|---|---|---|---|---|---|
| **SCHZ** | **N = 45** | | | | | |
| | **PST** | **0.953** | 0.63 | **~95%** | 0.862–1.000 | **< 0.0001** |
| | **SAT** | **0.863** | 1.29 | **>99%** | 0.728–0.999 | 0.0005 |
| **BIP** | **N = 36** | | | | | |
| | **PST** | **0.648** | −0.60 | **~85%** | 0.476–0.819 | ns |
| | **SAT** | **0.973** | **2.73** | **>99%** | 0.925–1.000 | **< 0.0001** |
| **FEP** | **N = 39 (38 for SAT)** | | | | | |
| | **PST** | **0.978** | **2.86** | **>99%** | 0.941–1.000 | **< 0.0001** |
| | **SAT** | **0.871** | 1.46 | **>99%** | 0.729–1.000 | 0.0003 |

***Note.*** AUC = area under the ROC curve; *d* = Cohen's effect size; CI = confidence interval; PST = Pictorial Size Test; SAT = Sound Appreciation Test; SCHZ = schizophrenia; BIP = bipolar disorder; FEP = first-episode psychosis. Values in **red** highlight Cohen's $d \geq 2.0$ or Power >99% or $p < 0.0001$. ns = not significant.

#### 2.2 Sound Appreciation Test (SAT)

The sixteen sounds of SAT in an experimental design of subjective magnitude can be found at: http://fonae.org/audios (last accessed [date]).

These sounds are hosted on the site of the National Forum on Schizophrenia, second edition May 21–23, 2025, held every two years.

*Note: **There are 20 sounds in total. We report only on the 16 pure-tone frequency sweeps**.*

### 2.3 Supplementary Figure 1.

PST (left) and SAT (right) means for each experimental group: SCHZ (top), BIP (middle), and FEP (bottom). Error bars are 95% confidence intervals. Significant differences are illustrated.

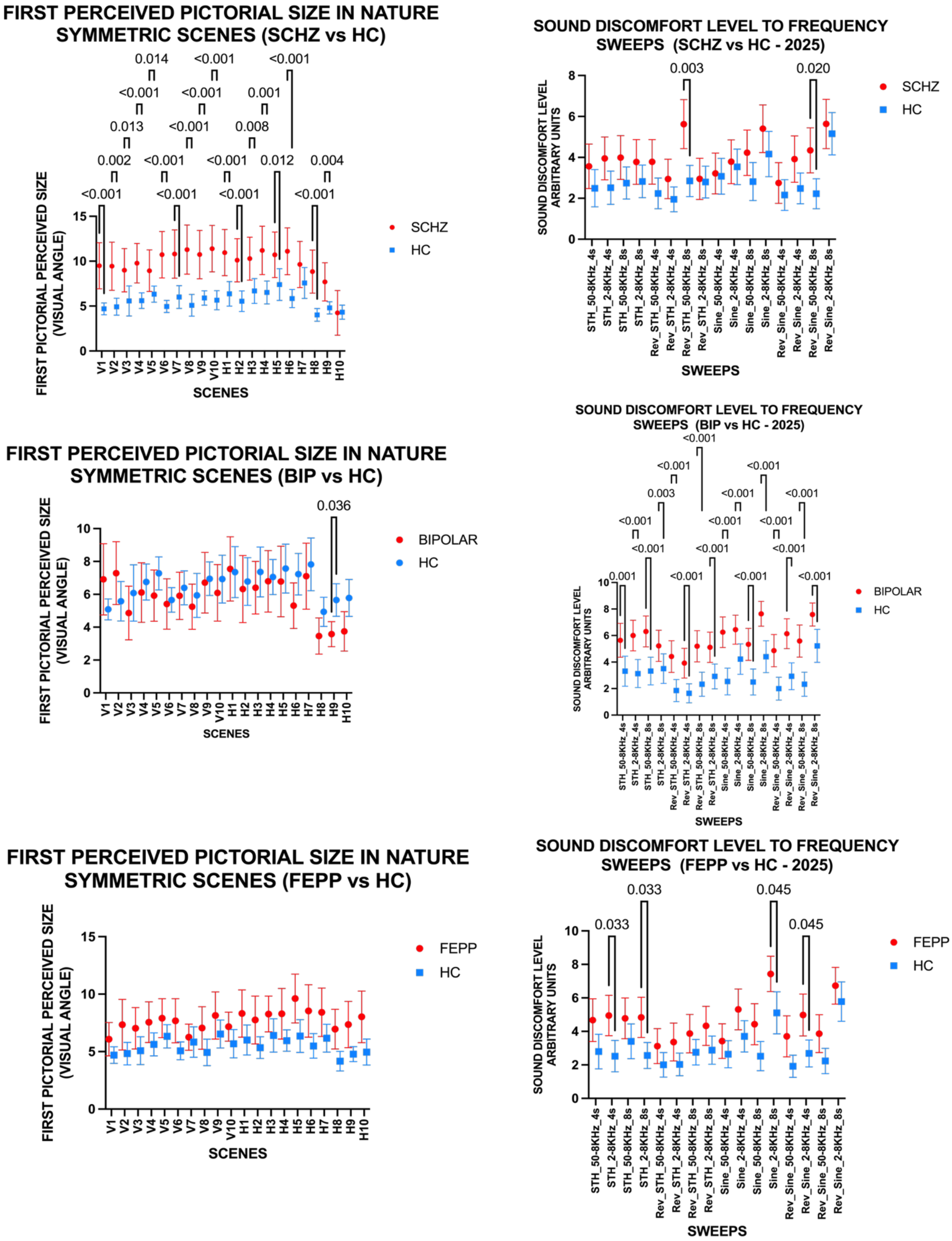

*[Insert*

**2.4 Supplementary Figure 2.**

PST means for perceived pictorial sizes per symmetry direction (vertical vs. horizontal) for SCHZ and BIP groups. Error bars are 95% confidence intervals.

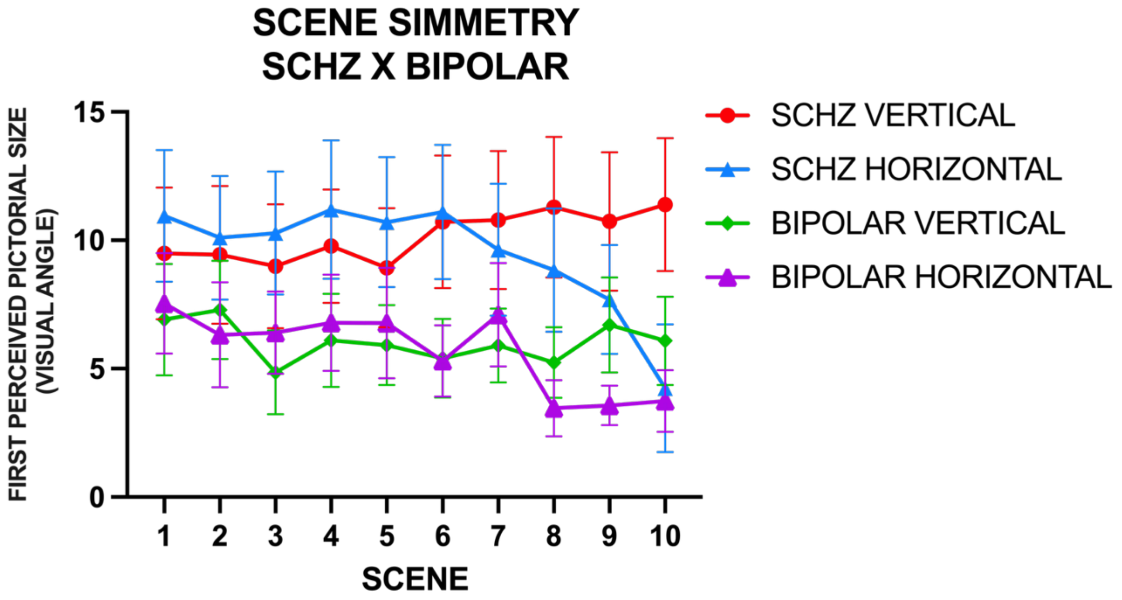

**2.5 Supplementary Figure 3.**

PST means across symmetry conditions comparing SCHZ vs. HC_SCHZ and FEP vs. HC_FEP.

**FEPP VS HC - 2025**

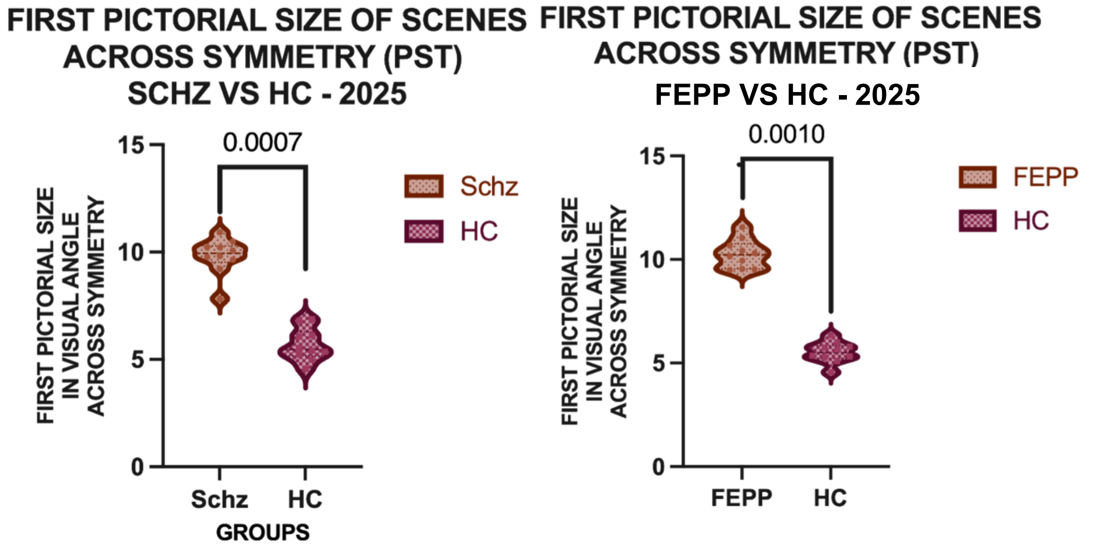

**2.6**

**Supplementary Figure 4**

PST responses to vertical and horizontal symmetric scenes for comparisons between SCHZ and BIP groups. These may be used for potential differential diagnosis. Error bars are 95% confidence intervals.

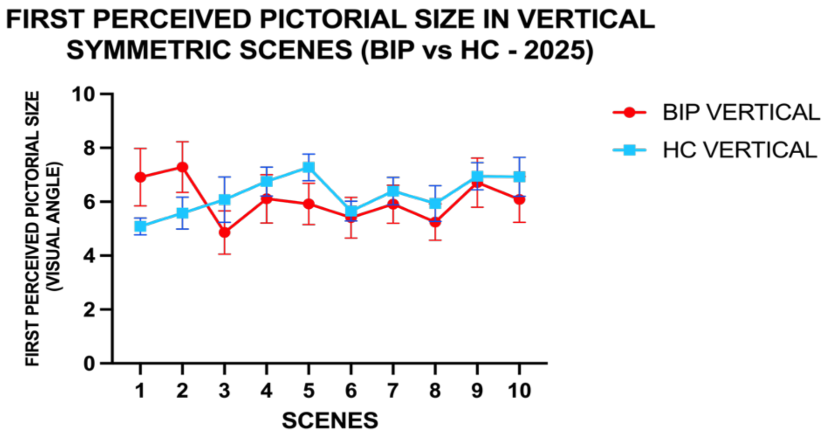

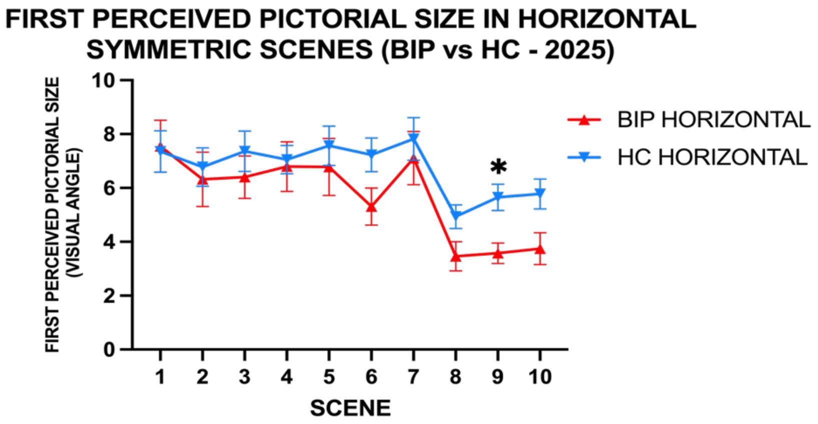

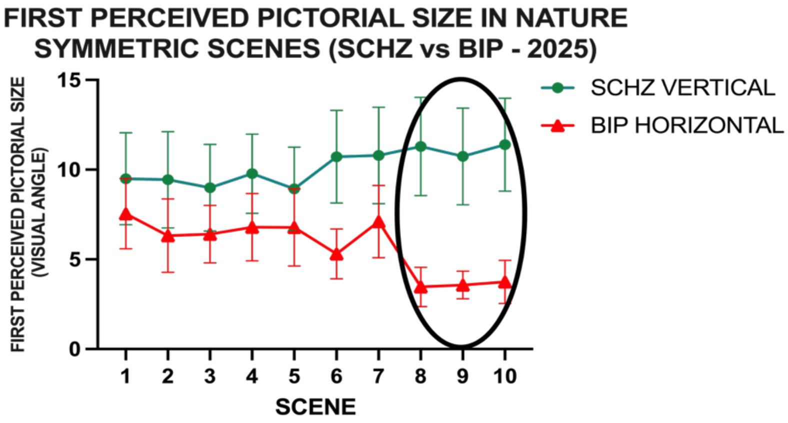

#### 2.7 Differential classification between SCHZ, BIP and undetermined

**Supplementary Table 3.** Empirically generated rules and their applications.

*PST = Pictorial Size Test score (degrees of visual angle, dva). SAT = Sound Appreciation Test score (0–10 sound discomfort level scale – arbitrary units).*

| RULE | Description | Prediction | Total | Correct | % correct |
|---|---|---|---|---|---|
| **R0** | PST ≤ 5 *dva* AND SAT ≥ 5 | *BIP* | 19 | 15 | 78.9% |
| **R1** | PST ≥ 10 *dva* | *SCHZ* | 17 | 13 | 76.5% |
| **R2** | PST > 5 *dva* AND SAT < 3 | *SCHZ* | 7 | 5 | 71.4% |
| **R3** | PST > 6 *dva* | *SCHZ* | 17 | 11 | 64.7% |
| **R4** | PST ≥ 4 *dva* AND (PST > 4.5 *dva* AND SAT ≤ 9) | *SCHZ* | 9 | 7 | 77.8% |
| **R5** | PST ≥ 4 *dva* AND SAT ∈ [PST±2.5] | *SCHZ* | 3 | 0 | 0.0% |
| **R6** | PST < 4 *dva* AND SAT > 5 | *BIP* | 0 | — | — |
| **R7** | UNDETERMINED | — | 9 | — | — |

**Supplementary Table 4.** Comparison after last rules adjustment: anterior SoundPics vs SoundPics V3.

| Metrics | Anterior (R4 corrected) | V3 (with R0) | Difference |
|---|---|---|---|
| Cases (determined) | 70 | 72 | +2 |
| Accuracy with clinical | 65.7% | 70.8% | +5.1% |
| Sensitivity BIP | 30.0% | 46.9% | +16.9% |
| Sensitivity SCHZ | 92.5% | 90.0% | −2.5% |
| Agreement with LDA | 77.1% | 84.7% | +7.6% |
| Undetermined | 11 | 9 | −2 |

**Supplementary Table 5.** Comparison with linear discriminant analysis (LDA) considering only the results of SCHZ ($N = 45$) and BIP ($N = 36$) groups.

| COMPARATIVE SUMMARY – LDA vs SoundPics V3 (with new RULE R0) | LDA | SoundPics V3 |
|---|---|---|
| **METRICS** | | |
| Total # cases | 81 | 81 |
| Cases categorized | 81 | 72 |
| Cases correct | 54 | 51 |
| Accuracy (clinic diagnosis coincidence) | 66.7% | 70.8% |
| Undetermined | 0 | 9 |

| **COMPARATIVE SUMMARY – LDA vs SoundPics V3 (with new RULE R0)** | **LDA** | **SoundPics V3** |
|---|---|---|
| % undetermined | 0% | 11.1% |
| **SENSITIVITIES** | | |
| Sensitivity BIP | — | 46.9% |
| Sensitivity SCHZ | — | 90.0% |
| **AGREEMENT BETWEEN METHODS** | | |
| Cases (agree) | 61 | 84.7% |
| Cases (disagree) | 11 | 15.3% |
| **CUT-OFF POINTS LDA** | | |
| PST isolated | 5.0 *dva* | — |
| SAT isolated | 5.6 (smaller → SCHZ) | — |
| Formula combined | $Z = 0.089 \times PST - 0.271 \times SAT + 0.862$ | — |
| Interpretation | $Z > 0$ → SCHZ; $Z < 0$ → BIP | — |
| AUC-ROC | 0.737 | — |

### 2.9 Supplementary Figure 5.

Comparative algorithm: LDA vs SoundPics V3. **Left panel:** ROC curves for individual predictors (PST, SAT) and combined LDA (*n* = 81). **Right panel:** ROC curves for SoundPics V3 (*n* = 72 determined cases) and LDA (*n* = 81). AUC values are shown in the table below. **Table:** Comparative summary of accuracy, AUC-ROC, sensitivities, agreement with LDA, and statistical power.

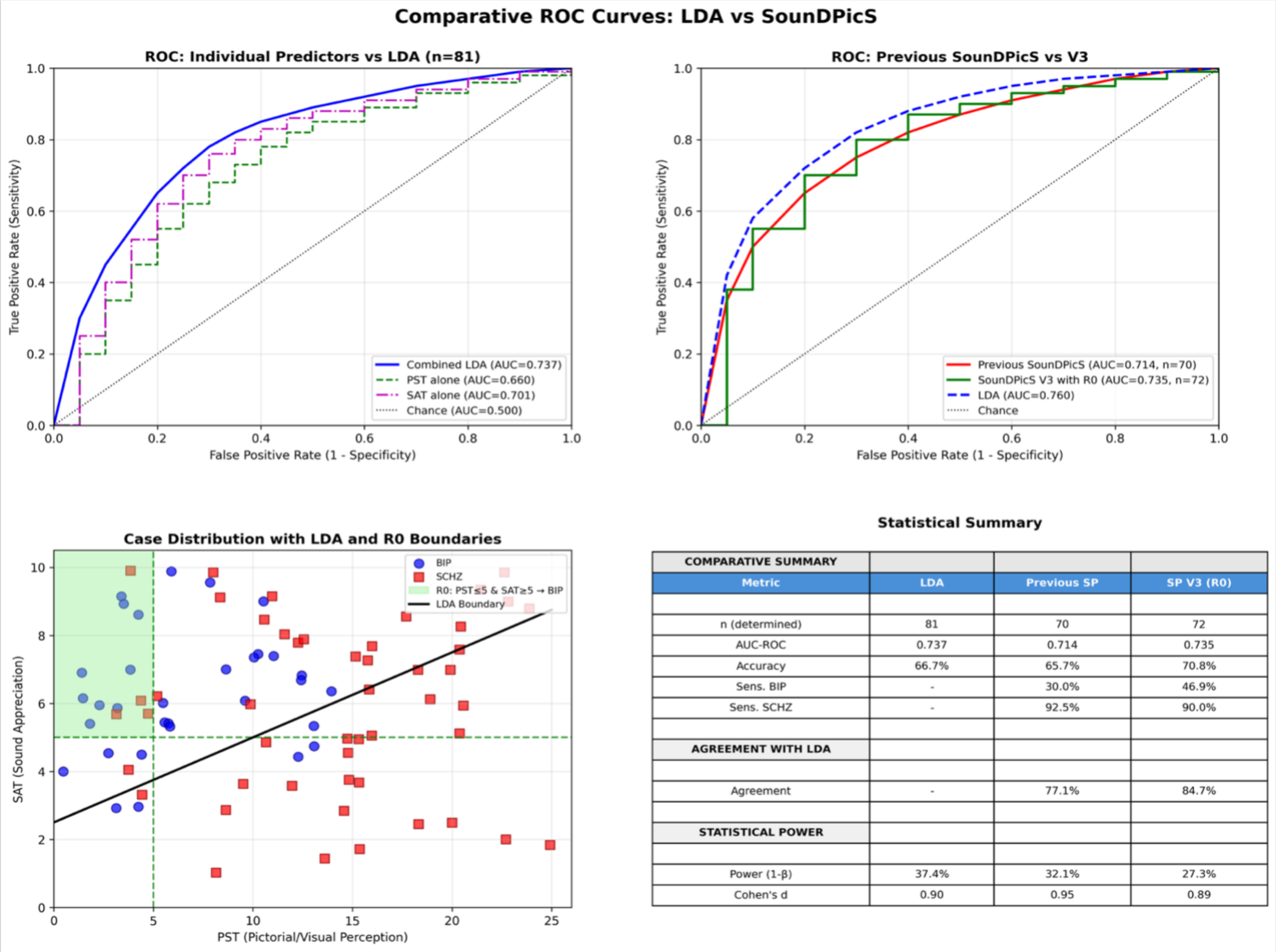

| COMPARATIVE SUMMARY | | | |
|---|---|---|---|
| Metric | LDA | Previous SP | SP V3 (R0) |
| n (determined) | 81 | 70 | 72 |
| AUC-ROC | 0.737 | 0.714 | 0.735 |
| Accuracy | 66.7% | 65.7% | 70.8% |
| Sens. BIP | - | 30.0% | 46.9% |
| Sens. SCHZ | - | 92.5% | 90.0% |
| AGREEMENT WITH LDA | | | |
| Agreement | - | 77.1% | 84.7% |
| STATISTICAL POWER | | | |
| Power (1-β) | 37.4% | 32.1% | 27.3% |
| Cohen's d | 0.90 | 0.95 | 0.89 |

*Formatting assistance for this figure was provided by Claude AI tool; all underlying data and statistical analyses are original and the responsibility of the authors.*

**END OF SUPPLEMENTARY MATERIAL**
